# Risk for Transportation of 2019 Novel Coronavirus (COVID-19) from Wuhan to Cities in China

**DOI:** 10.1101/2020.01.28.20019299

**Authors:** Zhanwei Du, Lin Wang, Simon Cauchemez, Xiaoke Xu, Xianwen Wang, Benjamin J. Cowling, Lauren Ancel Meyers

## Abstract

On January 23, 2020, China quarantined Wuhan to contain an emerging coronavirus (COVID-19). We estimated the probability of transportation of COVID-19 from Wuhan to 369 cities in China before the quarantine. The expected risk is >50% in 130 (95% CI 89–190) cities and >99% in the 4 largest metropolitan areas of China.

---

In December 2019, a novel coronavirus (COVID-19) emerged in Wuhan, China (1). On January 30, 2020, the World Health Organization (WHO) declared the outbreak a public health emergency of international concern (2). By January 31, 2020, 192 fatalities and 3,215 laboratory-confirmed cases were reported in Wuhan; 8,576 additional cases were spread across >300 cities in mainland China; and 127 exported cases were reported in 23 countries/states spanning Asia, Europe, Oceania, and North America. The rapid global expansion, rising fatalities, unknown animal reservoir, and evidence of person-to-person transmission potential (3,8) initially resembled the 2003 SARS epidemic and raised concerns about global spread.

On January 22, 2020, China announced a travel quarantine of Wuhan and by January 30, expanded the radius to include 16 cities, encompassing a population of 45 million. At the time of the quarantine, China was already 2 weeks into the 40-day Spring Festival, during which several billion people travel throughout China to celebrate the Lunar New Year (4). Considering the timing of exported COVID-19 cases reported outside of China, we estimate that only 8.95% (95% CrI 2.22% - 28.72%) of cases infected in Wuhan by January 12 might have been confirmed by January 22, 2020. By limiting our estimate to infections occurring ≥10 days before the quarantine, we account for an estimated 5–6-day incubation period and 4–5 days between symptom onset and case detection (3,5,6,8) (Appendix). The low detection rate coupled with an average lag of 10 days between infection and detection (6) suggest that newly infected persons who traveled out of Wuhan just before the quarantine might have remained infectious and undetected in dozens of cities in China for days to weeks. Moreover, these silent importations already might have seeded sustained outbreaks that were not immediately apparent.

We estimated the probability of transportation of infectious COVID-19 cases from Wuhan to cities throughout China before January 23 by using a simple model of exponential growth coupled with a stochastic model of human mobility among 369 cities in China (Appendix). Given that an estimated 98% of all trips between Wuhan and other Chinese cities during this period were taken by train or car, our analysis of air, rail, and road travel data yields more granular risk estimates than possible with air passenger data alone (7).

By fitting our epidemiologic model to data on the first 19 cases reported outside of China, we estimate an epidemic doubling time of 7.31 days (95% CrI 6.26 - 9.66 days) and a cumulative total of 12,400 (95% CrI 3,112–58,465) infections in Wuhan by January 22, 2020 (Appendix). Both estimates are consistent with a recent epidemiologic analysis of the first 425 cases confirmed in Wuhan (8). By assuming these rates of early epidemic growth, we estimate that 130 cities in China have ≥50% chance of having a COVID-19 case imported from Wuhan in the 3 weeks preceding the quarantine (Figure). By January 26th, 107 of these high-risk cities had reported cases and 23 had not, including 5 cities with importation probabilities >99% and populations >2 million: Bazhong, Fushun, Laibin, Ziyang, and Chuxiong. Under our lower bound estimate of 6.26 days for the doubling time, 190/369 cities lie above the 50% threshold for importation. Our risk assessment identified several cities throughout China likely to be harboring yet undetected cases of COVID-19 a week after the quarantine, suggesting that early 2020 ground and rail travel seeded cases far beyond the Wuhan region under quarantine.

Our conclusions are based on several key assumptions. To design our mobility model, we used data from Tencent (https://heat.qq.com), a major social media company that hosts applications, including WeChat (≈1.13 billion active users in 2019) and QQ (≈808 million active users in 2019) (9); consequently, our model might be demographically biased by the Tencent user base. Further, considerable uncertainty regarding the lag between infection and case detection remains. Our assumption of a 10-day lag is based on early estimates for the incubation period of COVID-19 (8) and prior estimates of the lag between symptom onset and detection for SARS (10). We expect that estimates for the doubling time and incidence of COVID-19 will improve as reconstructed linelists and more granular epidemiologic data become available (Appendix). However, our key qualitative insights likely are robust to these uncertainties, including extensive pre-quarantine COVID-19 exportations throughout China and far greater case counts in Wuhan than reported before the quarantine.

**Figure 1.**
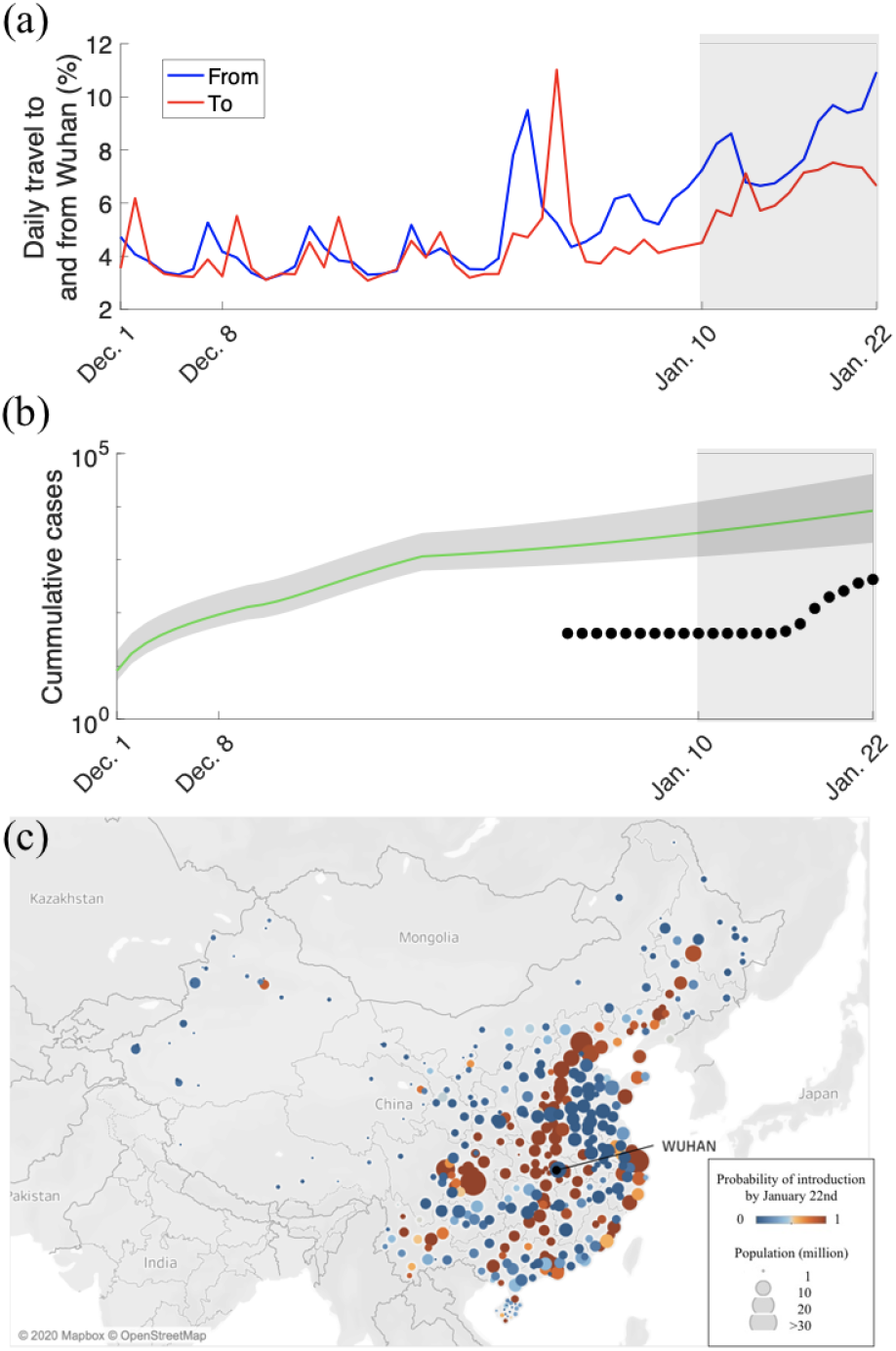
Risks for COVID-19 transportation from Wuhan, China before a quarantine was imposed on January 23, 2020. a) Daily travel volume to and from Wuhan, given as a percentage of the Wuhan population. The shading on the right indicates the start of Spring Festival season on January 10, 2020, which is a peak travel period in China. b) Estimated and reported daily prevalence of COVID-19 in Wuhan. The green line and shading indicate model estimates of cumulative cases since December 1, 2019 with 95% CrI bounds, based on an epidemic doubling time of 7.38 days (95% CrI 5.58–8.92 days). Black points are cumulative confirmed case counts during January 1–22, 2020 (*11*). The shading on the right indicates the start of Spring Festival season. c) Map generated by using Mapbox (https://www.mapbox.com) representing the probability that ≥1 COVID-19 case infected in Wuhan traveled to cities in China by January 22, 2020. The 131 cities above a risk threshold of 50% are indicated in orange; the 239 cities below the threshold are indicated in blue.

## Data Availability

Data will become available upon publication in a peer-reviewed journal.

## Supplementary Appendix

### Data

We analyzed the daily number of passengers traveling between Wuhan and 369 other cities in mainland China. We obtained mobility data from the location-based services of Tencent (https://heat.qq.com). Users permit Tencent to collect their real-time location information when they install applications, such as WeChat (≈1.13 billion active users in 2019) and QQ (≈808 million active users in 2019), and Tencent Map. By using the geolocation of users over time, Tencent reconstructed anonymized origin–destination mobility matrices by mode of transportation (air, road, and train) between 370 cities in China, including 368 cities in mainland China and the Special Administrative Regions of Hong Kong and Macau. The data are anonymized and include 28 million trips to and 32 million trips from Wuhan, during December 3, 2016–January 24, 2017. We estimated daily travel volume during the 7 weeks preceding the Wuhan quarantine, December 1, 2019–January 22, 2020, by aligning the dates of the Lunar New Year, resulting in a 3-day shift. To infer the number of new infections in Wuhan per day during December 1, 2019–January 22, 2020, we used the mean daily number of passengers traveling to the top 27 foreign destinations from Wuhan during 2018–2019, which were provided in other recent studies (*1*–*3*).

### Model

We considered a simple hierarchical model to describe the dynamics of 2019 novel coronavirus (COVID-19) infections, detections, and spread.

### Epidemiologic Model

By using epidemiologic evidence from the first 425 cases of COVID-19 confirmed in Wuhan by January 22, 2020 (*4*), we made the following assumptions regarding the number of new cases, *dI*_*w*_(*t*), infected in Wuhan per day, *t*.

- The COVID-19 epidemic was growing exponentially during December 1, 2019–January 22, 2020, as determined by the following:

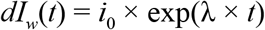

in which *i*_0_ denotes the number of initial cases on December 1, 2019 (*5*), and λ denotes the epidemic growth rate during December 1, 2019–January 22, 2020.
- After infection, new cases were detected with a delay of *D* = 10 days (*6*), which comprises an incubation period of 5–6 days (*4,7*–*11*) and a delay from symptom onset to detection of 4–5 days (*12,13*). During this 10-day interval, we labeled cases as infected. Given the uncertainty in these estimates, we also performed the estimates by assuming a shorter delay (*D* = 6 days) and a longer delay (*D* = 14 days) between infection and case detection (Appendix Table 2).

Our model can be improved by incorporating the probability distribution for the delay between infection and detection, as reconstructed linelists (*14*–*17*) and more granular epidemiologic data are becoming available.

Under these assumptions, we calculated the number of infectious cases at time, *t*, by the following:

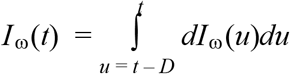

The prevalence of infectious cases is given by the following:

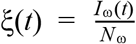

in which *N* ω = 11.08 million, the population of Wuhan.

### Mobility Model

We assume that visitors to Wuhan have the same daily risk for infection as residents of Wuhan and construct a nonhomogeneous Poisson process model (*18*–*20*) to estimate the exportation of COVID-19 by residents of and travelers to Wuhan. Let *W*_*j,t*_ denote the number of Wuhan residents that travel to city *j* at time *t*, and *M*_*j,t*_ denote the number of travelers from city *j* to Wuhan at time *t*. Then, the rate at which infected residents of Wuhan travel to city *j* at time *t* is given γ_*j,t*_ = ξ(*t*) × *W*_*j,t*_, and the rate at which travelers from city *j* get infected in Wuhan and return to their home city while still infected is Ψ_*j,t*_ = ξ(*t*) × *M*_*j,t*_. This model assumes that newly infected visitors to Wuhan will return to their home city while still infectious.

Based on this model, the probability of introducing ≥1 cases from Wuhan to city *j* by time *t* is given by

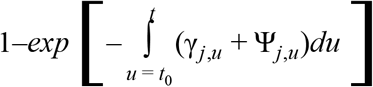

where *t*_0_ denotes the beginning of the study period, December 1, 2019.

### Inference of Epidemic Parameters

We applied a likelihood-based method to estimate our model parameters, including the number of initial cases *i*_0_ and the epidemic growth rate λ, from the arrival times of the 19 reported cases transported from Wuhan to 11 cities outside of China, as of January 22, 2020 (Appendix Table 1). All 19 cases were Wuhan residents. We aggregated all other cities without cases reported by January 22, 2020 into a single location (*j* = 0).

Let *N*_*j*_ denote the number of infected Wuhan residents who were detected in location *j* outside of China, <u>denote the time at which</u> the *i*-th Wuhan resident case was detected in location *j, χ*_*j*,0_ denote the time at which international surveillance for infected travelers from Wuhan began (January 1, 2020) (*21*), and *E* denote the end of the study period (January 22, 2020). As indicated above, the rate at which infected residents of Wuhan arrive at location *j* at time *t* is γ_*j,t*_. Then, the likelihood for all 19 cases reported outside of China by January 22, 2020 is given by

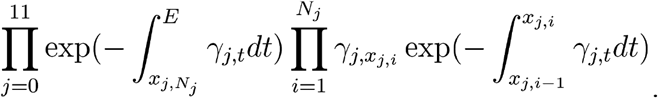

which yields the following log-likelihood function:

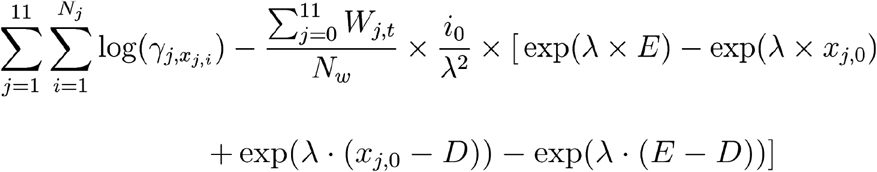

### Parameter Estimation

We directly estimated the number of initial cases, *i*_0_, on December 1, 2019, and the epidemic growth rate, λ, during December 1, 2019–January 22, 2020. We infer the epidemic parameters in a Bayesian framework by using the Markov Chain Monte Carlo (MCMC) method with Hamiltonian Monte Carlo sampling and noninformative flat prior. From these, we derive the doubling time of incident cases as *d*_*T*_ = log(2)/λ and the cumulative number of cases and of reported cases by January 22, 2020. We also derived the basic reproduction number by assuming a susceptible-exposed-infectious-recovery (SEIR) model for COVID-19, in which the incubation period is exponentially distributed with mean *L* in the range of 3 - 6 days and the infectious period is also exponentially distributed with mean *Z* in the range of 2 to 7 days. The reproduction number is then given by *R*_0_ = (1 + λ × *L*) × (1 + λ × *Z*).

We estimated the case detection rate in Wuhan by taking the ratio between the number of reported cases in Wuhan by January 22, 2020 and our estimates for the number of infections occurring ≥10 days prior (i.e., by January 12, 2020). We truncated our estimate 10 days before the quarantine to account for the estimated time between infection and case detection, assuming a 5–6 day incubation period (*4,7*–*11*) followed by 4–5 days between symptom onset and case detection (*12,13*). Given the uncertainty in these estimates, we also provide estimates assuming shorter and longer delays in the lag between infection and case reporting (Appendix Table 3).

We ran 10 chains in parallel. Trace plot and diagnosis confirmed the convergence of MCMC chains with posterior median and 95% CrI estimates as follows:

- Epidemic growth rate, λ: 0.095 (0.072 - 0.111), corresponding to an epidemic doubling time of incident cases of 7.31 (95% CrI 6.26 - 9.66) days;
- Number of initial cases in Wuhan on December 1, 2019: 7.78 (95% CrI 5.09 - 18.27);
- Basic reproductive number, *R*_0_: 1.90 (95% CrI 1.47 - 2.59);
- Cumulative number of infections in Wuhan by January 22, 2020: 12,400 (95% CrI 3,112–58,465);
- Case detection rate by January 22, 2020: 8.95% (95% CrI 2.22% - 28.72%). This represents the ratio between the 425 confirmed cases in Wuhan during this period (*22*) and our estimate that 4,747 (95% CrI 1,480–19,151) cumulative infections occurred by January 12, 2020 (i.e., ≥10 days before the quarantine to account for the typical lag between infection and case detection).

## Acknowledgements

We thank Henrik Salje, Dongsheng Luo, Bo Xu, Cécile Tran Kiem, Dong Xun, and Lanfang Hu for helpful discussions. We acknowledge the financial support from NIH (U01 GM087719), the Investissement d’Avenir program, the Laboratoire d’Excellence Integrative Biology of Emerging Infectious Diseases program (Grant ANR-10-LABX-62-IBEID), European Union V.E.O project, the Open Fund of Key Laboratory of Urban Land Resources Monitoring and Simulation, Ministry of Land and Resources (KF-2019-04-034), and the National Natural Science Foundation of China (61773091).

Code for estimating epidemiological parameters and probabilities of case introductions, as well as aggregate mobility data are available at: https://github.com/linwangidd/2019nCoV_EID. Aggregate data are also available in Appendix Table S3. Additional code and data requests should be addressed to Lauren Ancel Meyers (, 512-471-4950).

---

**Figure S1.**
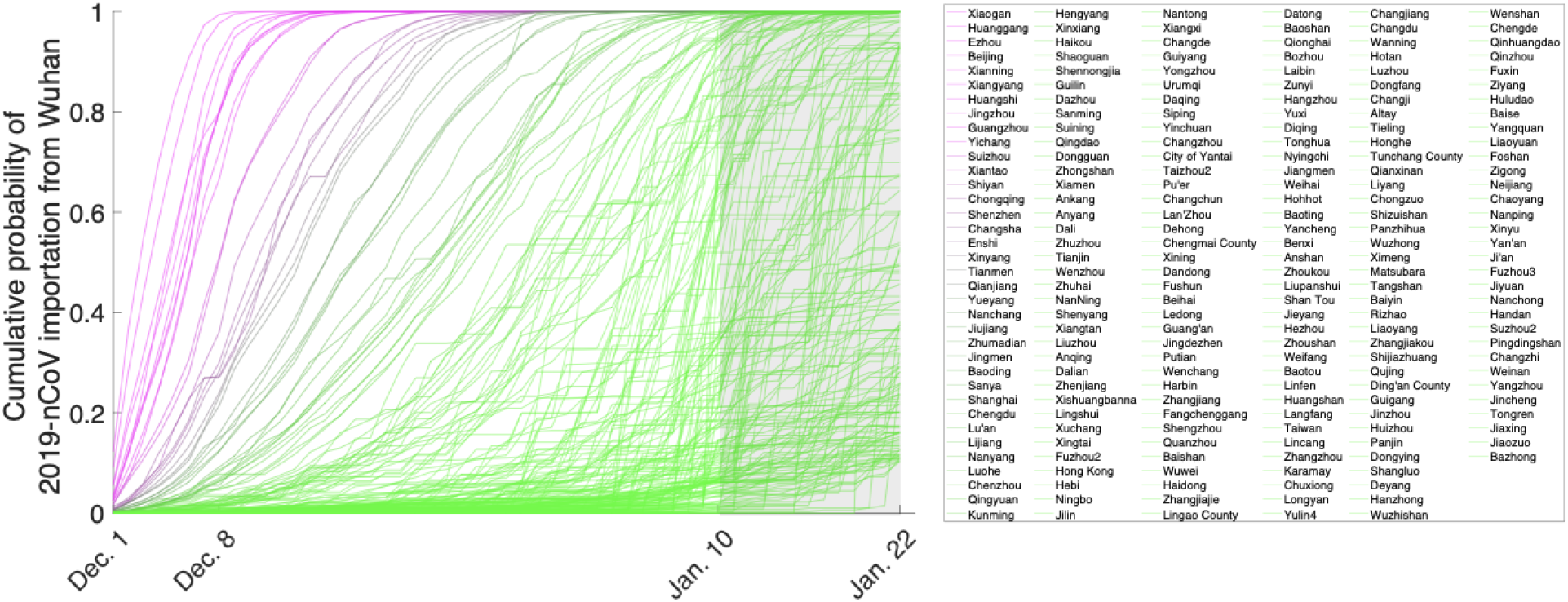
The risk for introduction of 2019 novel coronavirus (COVID-19) from Wuhan to other cities in China before the January 23, 2020 quarantine of Wuhan. Lines indicate probabilities that at ≥1 person infected with COVID-19 in Wuhan arrived in a listed city by the date indicated on the x-axis. The estimates were calculated by using mobility data collected from the location-based services of Tencent (https://heat.qq.com) during December 10, 2017–January 24, 2018, the timeframe that corresponds to the Spring Festival travel period of December 8, 2019–January 22, 2020. All cities with an expected importation probability >10% by January 22, 2020 (n = 212) are shown.

**Figure S2.**
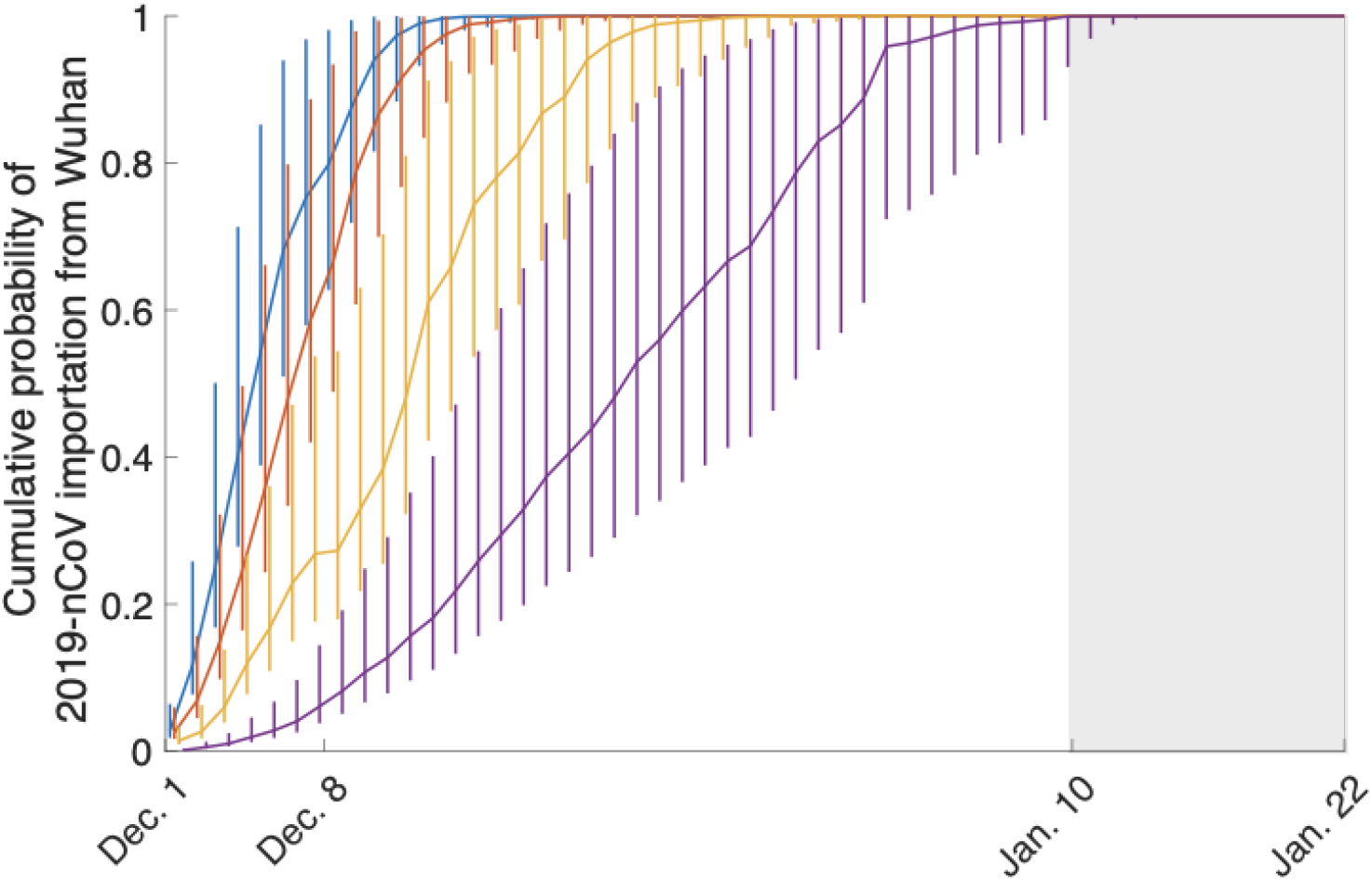
Uncertainty analysis representing the number of 2019 novel coronavirus (COVID-19) exposures in Wuhan per day. Lines show the probability that ≥1 transportation of COVID-19 infection occurred from Wuhan to Beijing, Guangzhou, Shenzhen, and Shanghai during December 8, 2020–January 22, 2020. Error bars indicate 95% credible intervals.

**Figure S3.**
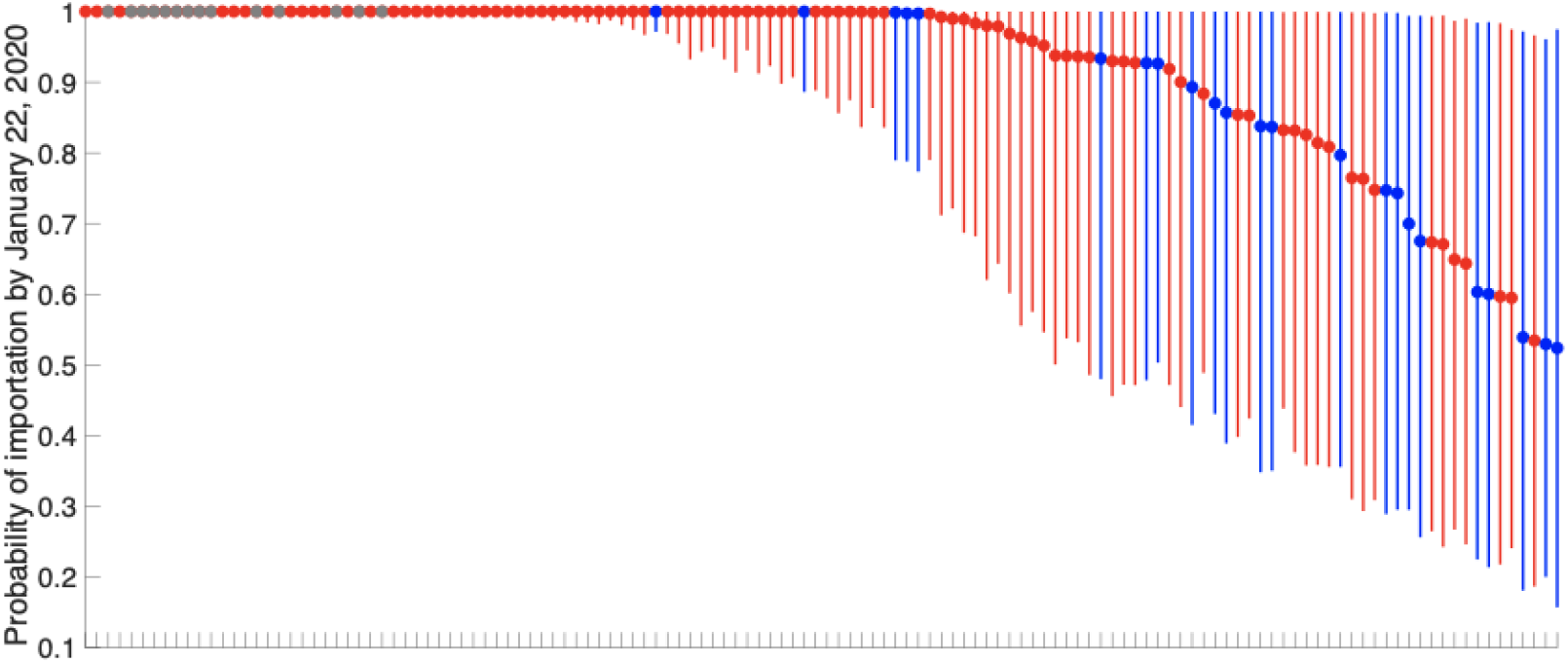
Risk for transportation of 2019 novel coronavirus (COVID-19) from Wuhan to 130 cities in China by January 23, 2020. All cities represented have mean importation probability >50%. As of January 26, 2020, 82.3% (107/130) of these cities had reported cases. Grey circles indicate cities that were included in the quarantine as of January 24, 2020. Red circles indicate cities outside the quarantine area with confirmed cases; blue circles indicate cities outside the quarantine area without confirmed cases as of January 26th, 2020.

**Table S1.** Cases of 2019 novel coronavirus detected outside of China*

| Country/State | City | Date, 2020 |
| --- | --- | --- |
| Thailand | Bangkok | Jan 8 |
| Thailand | Bangkok | Jan 17 |
| Thailand | Bangkok | Jan 19 |
| Thailand | Bangkok | Jan 21 |
| Thailand | Chiang Mai | Jan 21 |
| Nepal | Kathmandu | Jan 9 |
| Vietnam | Hanoi | Jan 13 |
| United States | Chicago | Jan 13 |
| United States | Seattle | Jan 15 |
| Singapore |  | Jan 21 |
| Korea | Seoul | Jan 19 |
| Korea | Seoul | Jan 22 |
| Japan | Tokyo | Jan 18 |
| Japan | Tokyo | Jan 19 |
| Taiwan | Taipei | Jan 20 |
| Taiwan | Taipei | Jan 21 |
| Taiwan | Taipei | Jan 21 |
| Australia | Sydney | Jan 18 |
| Australia | Sydney | Jan 20 |

\*As of January 22, 2020.

**Table S2.** Sensitivity analysis for the delay between infection and case confirmation, assuming that cases were confirmed either 6 d, 10 d (baseline), or 14 d after infection

| Delay ($D$) from infection to case reporting | Posterior median (95% CrI) |
| --- | --- |
| $D = 6$ d | |
| Epidemic doubling time, d | 6.79 (5.88 - 8.64) |
| Initial number of cases on December 1, 2019, $i_0$ | 7.95 (5.10 - 18.43) |
| Basic reproduction number, $R_0$ | 1.98 (1.54 - 2.71) |
| Cumulative cases by January 22, 2020 | 17,376 (4,410 - 80,915) |
| Cumulative cases by January 16, 2020 ($D = 6$ d before January 22, 2020) | 9,362 (2,696 - 39,705) |
| Reporting rate through January 22, 2020 | 4.54% (1.07% - 15.8%) |
| $D = 10$ d | |
| Epidemic doubling time, d | 7.31 (6.26 - 9.66) |
| Initial number of cases on December 1, 2019, $i_0$ | 7.78 (5.09 - 18.27) |
| Basic reproduction number, $R_0$ | 1.90 (1.47 - 2.59) |
| Cumulative cases by January 22, 2020 | 12,400 (3,112–58,465) |
| Cumulative cases by January 16, 2020 ($D = 6$ d before January 22, 2020) | 4,747 (1,480–19,151) |
| Reporting rate through January 22, 2020 | 8.95% (2.22% - 28.72%) |
| $D = 14$ d | |
| Epidemic doubling time, d | 7.64 (6.49 - 10.36) |
| Initial number of cases on December 1, 2019, $i_0$ | 7.62 (5.09 - 18.13) |
| Basic reproduction number, $R_0$ | 1.86 (1.44 - 2.52) |
| Cumulative cases by January 22, 2020 | 10,229 (2,564 - 48,681) |
| Cumulative cases by January 16, 2020 ($D = 6$ d before January 22, 2020) | 2,805 (957 - 10,758) |
| Reporting rate through January 22, 2020 | 15.15% (3.95% - 44.41%) |

**Table S3.** Mobility between Wuhan and 369 cities in China during December 3, 2016–January 24, 2017*

| ID | City | Total trips | From Wuhan | To Wuhan | 2016 population, millions |
| --- | --- | --- | --- | --- | --- |
| 1 | Xiaogan | 9,646,286 | 5,333,682 | 4,312,604 | 4.90 |
| 2 | Huanggang | 7,786,732 | 4,436,928 | 3,349,804 | 6.32 |
| 3 | Xianning | 3,987,334 | 2,149,524 | 1,837,810 | 2.53 |
| 4 | Ezhou | 3,921,153 | 1,956,195 | 1,964,958 | 1.07 |
| 5 | Beijing | 3,858,883 | 1,508,938 | 2,349,945 | 21.73 |
| 6 | Jingzhou | 3,439,123 | 2,216,479 | 1,222,644 | 5.70 |
| 7 | Xiangyang | 3,160,473 | 1,959,413 | 1,201,060 | 5.64 |
| 8 | Huangshi | 2,787,922 | 1,521,685 | 1,266,237 | 2.47 |
| 9 | Guangzhou | 2,555,286 | 705,205 | 1,850,081 | 14.04 |
| 10 | Yichang | 2,266,974 | 1,420,349 | 846,625 | 4.13 |
| 11 | Shenzhen | 1,675,478 | 188,316 | 1,487,162 | 11.91 |
| 12 | Suizhou | 1,536,742 | 934,564 | 602,178 | 2.20 |
| 13 | Xiantao | 1,492,596 | 856,578 | 636,018 | 1.15 |
| 14 | Shiyan | 1,252,190 | 897,666 | 354,524 | 3.41 |
| 15 | Chongqing | 1,177,096 | 720,442 | 456,654 | 30.48 |
| 16 | Enshi | 869,910 | 610,937 | 258,973 | 4.56 |
| 17 | Tianmen | 716,794 | 447,408 | 269,386 | 1.29 |
| 18 | Changsha | 644,273 | 318,784 | 325,489 | 7.65 |
| 19 | Shanghai | 571,458 | 72,150 | 499,308 | 24.20 |
| 20 | Xinyang | 564,841 | 338,180 | 226,661 | 6.44 |
| 21 | Qianjiang | 489,747 | 288,200 | 201,547 | 0.96 |
| 22 | Jingmen | 408,465 | 269,703 | 138,762 | 2.90 |
| 23 | Yueyang | 352,512 | 185,672 | 166,840 | 5.68 |
| 24 | Zhumadian | 316,181 | 214,425 | 101,756 | 6.99 |
| 25 | Nanchang | 301,903 | 123,239 | 178,664 | 5.37 |
| 26 | Jiujiang | 229,539 | 106,873 | 122,666 | 4.85 |
| 27 | Baoding | 205,124 | 126,334 | 78,790 | 11.63 |
| 28 | Nanyang | 173,653 | 127,666 | 45,987 | 10.07 |
| 29 | Hengyang | 155,591 | 32,443 | 123,148 | 7.29 |
| 30 | Luohe | 153,337 | 103,153 | 50,184 | 2.64 |
| 31 | Sanya | 151,726 | 29,147 | 122,579 | 0.75 |
| 32 | Lijiang | 121,669 | 33,825 | 87,844 | 1.29 |
| 33 | Dazhou | 120,983 | 120,983 | 0 | 5.60 |
| 34 | Luan | 117,242 | 53,698 | 63,544 | 4.77 |
| 35 | Qingyuan | 116,218 | 35,704 | 80,514 | 3.85 |
| 36 | Chengdu | 113,938 | 50,532 | 63,406 | 15.92 |
| 37 | Kunming | 108,452 | 46,613 | 61,839 | 6.73 |
| 38 | Chenzhou | 102,565 | 18,274 | 84,291 | 4.71 |
| 39 | Guilin | 100,723 | 92,078 | 8,645 | 5.01 |
| 40 | Shaoguan | 94,847 | 11,483 | 83,364 | 2.96 |
| 41 | Shijiazhuang | 93,102 | 70,128 | 22,974 | 10.78 |
| 42 | Ankang | 81,065 | 81,065 | 0 | 2.66 |
| 43 | Xinxiang | 73,246 | 54,707 | 18,539 | 5.74 |
| 44 | Shennongjia | 66,818 | 37,240 | 29,578 | 0.08 |
| 45 | Suining | 64,847 | 43,223 | 21,624 | 3.30 |
| 46 | Haikou | 64,774 | 30,848 | 33,926 | 2.24 |
| 47 | Shenyang | 64,258 | 33,663 | 30,595 | 8.29 |
| 48 | Hanzhong | 58,082 | 58,074 | 8 | 3.45 |
| 49 | Anyang | 57,825 | 38,146 | 19,679 | 5.13 |
| 50 | Dongguan | 57,672 | 44,125 | 13,547 | 8.26 |
| 51 | Liuzhou | 56,640 | 43,180 | 13,460 | 3.96 |
| 52 | Zhuzhou | 53,890 | 27,321 | 26,569 | 4.02 |
| 53 | Handan | 52,175 | 42,872 | 9,303 | 9.49 |
| 54 | Fuzhou2 | 50,264 | 11,069 | 39,195 | 7.57 |
| 55 | Sanming | 48,697 | 36,007 | 12,690 | 2.55 |
| 56 | NanNing | 47,505 | 33,242 | 14,263 | 7.06 |
| 57 | Xingtai | 44,627 | 33,727 | 10,900 | 7.32 |
| 58 | Xuchang | 44,397 | 41,839 | 2,558 | 4.38 |
| 59 | Anqing | 41,590 | 17,398 | 24,192 | 4.61 |
| 60 | Dali | 40,710 | 17,524 | 23,186 | 3.56 |
| 61 | Yongzhou | 40,530 | 40,530 | 0 | 5.47 |
| 62 | Xiamen | 40,039 | 14,993 | 25,046 | 3.92 |
| 63 | Qingdao | 36,803 | 21,919 | 14,884 | 9.20 |
| 64 | Nanchong | 33,778 | 33,764 | 14 | 6.40 |
| 65 | Pingdingshan | 30,833 | 25,945 | 4,888 | 4.98 |
| 66 | Tieling | 30,807 | 13,535 | 17,272 | 2.65 |
| 67 | Putian | 30,488 | 21,972 | 8,516 | 2.89 |
| 68 | Zhuhai | 30,263 | 20,698 | 9,565 | 1.68 |
| 69 | Wenzhou | 29,609 | 15,634 | 13,975 | 9.18 |
| 70 | Jiaozuo | 26,455 | 26,445 | 10 | 3.55 |
| 71 | Guangan | 25,597 | 24,288 | 1,309 | 3.26 |
| 72 | Nantong | 22,577 | 7,753 | 14,824 | 7.30 |
| 73 | Xiangtan | 22,283 | 7,879 | 14,404 | 2.84 |
| 74 | Langfang | 21,900 | 7,301 | 14,599 | 4.62 |
| 75 | Tianjin | 21,343 | 12,018 | 9,325 | 15.62 |
| 76 | Zhenjiang | 21,092 | 17,499 | 3,593 | 3.18 |
| 77 | Suzhou2 | 20,366 | 0 | 20,366 | 10.65 |
| 78 | Huludao | 19,114 | 18,044 | 1,070 | 2.55 |
| 79 | Jincheng | 18,326 | 18,318 | 8 | 2.32 |
| 80 | Siping | 17,782 | 3,610 | 14,172 | 3.20 |
| 81 | Dalian | 17,190 | 6,147 | 11,043 | 6.99 |
| 82 | Zhongshan | 17,181 | 14,989 | 2,192 | 3.23 |
| 83 | Shangluo | 17,033 | 16,740 | 293 | 2.37 |
| 84 | Beihai | 16,142 | 6,120 | 10,022 | 1.64 |
| 85 | Changzhi | 14,729 | 14,729 | 0 | 3.44 |
| 86 | Bazhong | 14,705 | 14,705 | 0 | 3.31 |
| 87 | Hebi | 14,173 | 9,224 | 4,949 | 1.61 |
| 88 | Xishuangbanna | 11,767 | 6,146 | 5,621 | 1.17 |
| 89 | Hong Kong | 11,453 | 5,823 | 5,630 | 7.45 |
| 90 | Zhoukou | 11,066 | 11,066 | 0 | 8.82 |
| 91 | Urumqi | 10,893 | 10,058 | 835 | 3.52 |
| 92 | Harbin | 10,110 | 5,991 | 4,119 | 10.98 |
| 93 | Ningbo | 9,964 | 5,272 | 4,692 | 7.88 |
| 94 | Weinan | 9,743 | 9,743 | 0 | 5.37 |
| 95 | Changchun | 9,379 | 6,040 | 3,339 | 7.51 |
| 96 | Laibin | 9,200 | 8,652 | 548 | 2.20 |
| 97 | Panjin | 9,130 | 8,398 | 732 | 1.44 |
| 98 | Xiangxi | 8,616 | 2,506 | 6,110 | 2.64 |
| 99 | City of Yantai | 8,223 | 4,390 | 3,833 | 7.06 |
| 100 | Yuxi | 7,895 | 5,513 | 2,382 | 2.38 |
| 101 | Tangshan | 7,604 | 7,152 | 452 | 7.84 |
| 102 | Lingshui | 7,477 | 1,792 | 5,685 | 0.36 |
| 103 | Xining | 7,414 | 5,460 | 1,954 | 2.33 |
| 104 | Liyang | 7,291 | 7,291 | 0 | 3.63 |
| 105 | Hezhou | 7,274 | 7,274 | 0 | 2.04 |
| 106 | Hangzhou | 7,112 | 797 | 6,315 | 9.19 |
| 107 | Nanping | 7,053 | 3,854 | 3,199 | 2.66 |
| 108 | Yinchuan | 6,789 | 3,364 | 3,425 | 2.08 |
| 109 | Changzhou | 6,761 | 6,761 | 0 | 4.71 |
| 110 | Zigong | 6,705 | 6,681 | 24 | 2.78 |
| 111 | Fushun | 6,576 | 5,816 | 760 | 2.07 |
| 112 | Puer | 6,335 | 3,781 | 2,554 | 2.62 |
| 113 | Taizhou2 | 6,269 | 2,362 | 3,907 | 6.08 |
| 114 | Changde | 6,131 | 4,946 | 1,185 | 5.84 |
| 115 | Jinzhou | 6,034 | 5,919 | 115 | 3.06 |
| 116 | Chengde | 5,937 | 5,786 | 151 | 3.53 |
| 117 | Yangzhou | 5,840 | 5,840 | 0 | 4.49 |
| 118 | Qijing | 5,396 | 5,041 | 355 | 6.08 |
| 119 | Yangquan | 5,313 | 5,269 | 44 | 1.40 |
| 120 | Anshan | 5,308 | 4,044 | 1,264 | 3.61 |
| 121 | Guiyang | 5,183 | 3,207 | 1,976 | 4.70 |
| 122 | Zhangjiajie | 5,157 | 4,112 | 1,045 | 1.53 |
| 123 | Quanzhou | 5,127 | 1,705 | 3,422 | 8.58 |
| 124 | Jian | 5,126 | 0 | 5,126 | 4.92 |
| 125 | Wuwei | 4,965 | 4,679 | 286 | 1.82 |
| 126 | Ledong | 4,807 | 3,014 | 1,793 | 0.53 |
| 127 | Liaoyang | 4,554 | 4,255 | 299 | 1.84 |
| 128 | Jiangmen | 4,550 | 4,439 | 111 | 4.54 |
| 129 | LanZhou | 4,154 | 2,226 | 1,928 | 3.71 |
| 130 | Qinhuangdao | 4,147 | 3,883 | 264 | 3.09 |
| 131 | Ziyang | 3,971 | 3,933 | 38 | 2.54 |
| 132 | Jingdezhen | 3,971 | 1,916 | 2,055 | 1.65 |
| 133 | Diqing | 3,933 | 1,123 | 2,810 | 0.41 |
| 134 | Shengzhou | 3,871 | 1,134 | 2,737 | 0.96 |
| 135 | Dehong | 3,645 | 1,735 | 1,910 | 1.29 |
| 136 | Panzhuhua | 3,536 | 2,197 | 1,339 | 1.24 |
| 137 | Neijiang | 3,526 | 3,493 | 33 | 3.75 |
| 138 | Foshan | 3,422 | 3,157 | 265 | 7.46 |
| 139 | Zhangjiang | 3,377 | 1,426 | 1,951 | 7.27 |
| 140 | Qionghai | 3,287 | 1,321 | 1,966 | 0.51 |
| 141 | Hohhot | 3,278 | 2,905 | 373 | 3.09 |
| 142 | Luzhou | 3,155 | 2,974 | 181 | 4.31 |
| 143 | Dandong | 3,136 | 2,165 | 971 | 2.41 |
| 144 | Deyang | 3,135 | 2,962 | 173 | 3.52 |
| 145 | Baoshan | 3,114 | 1,767 | 1,347 | 2.61 |
| 146 | Fangchenggang | 2,967 | 1,486 | 1,481 | 0.93 |
| 147 | Chuxiong | 2,966 | 2,419 | 547 | 2.74 |
| 148 | Datong | 2,881 | 1,914 | 967 | 3.42 |
| 149 | Zunyi | 2,775 | 1,544 | 1,231 | 6.23 |
| 150 | Jilin | 2,464 | 1,031 | 1,433 | 4.24 |
| 151 | Haidong | 2,421 | 1,062 | 1,359 | 1.45 |
| 152 | Baotou | 2,378 | 1,947 | 431 | 2.86 |
| 153 | Chengmai County | 2,301 | 905 | 1,396 | 0.59 |
| 154 | Huangshan | 2,226 | 959 | 1,267 | 1.38 |
| 155 | Benxi | 2,166 | 1,886 | 280 | 1.71 |
| 156 | Wenchang | 2,087 | 1,124 | 963 | 0.56 |
| 157 | Liupanshui | 2,086 | 589 | 1,497 | 2.91 |
| 158 | Lingao County | 2,085 | 1,349 | 736 | 0.52 |
| 159 | Daqing | 2,062 | 715 | 1,347 | 2.76 |
| 160 | Bozhou | 2,031 | 1,014 | 1,017 | 0.48 |
| 161 | Honghe | 1,960 | 1,262 | 698 | 4.68 |
| 162 | Lincang | 1,901 | 927 | 974 | 2.52 |
| 163 | Yancheng | 1,855 | 790 | 1,065 | 7.24 |
| 164 | Shan Tou | 1,847 | 786 | 1,061 | 5.58 |
| 165 | Fuzhou3 | 1,846 | 0 | 1,846 | 4.00 |
| 166 | Zhangjiakou | 1,845 | 1,743 | 102 | 4.43 |
| 167 | Yiyang | 1,820 | 1,365 | 455 | 4.43 |
| 168 | Dongying | 1,794 | 1,624 | 170 | 2.13 |
| 169 | Tonghua | 1,792 | 749 | 1,043 | 2.17 |
| 170 | Jieyang | 1,765 | 940 | 825 | 6.09 |
| 171 | Dongfang | 1,759 | 894 | 865 | 0.44 |
| 172 | Huizhou | 1,745 | 1,694 | 51 | 4.78 |
| 173 | Weihai | 1,744 | 677 | 1,067 | 2.82 |
| 174 | Wanning | 1,741 | 792 | 949 | 0.57 |
| 175 | Jiyuan | 1,555 | 1,461 | 94 | 0.73 |
| 176 | Longyan | 1,535 | 508 | 1,027 | 2.63 |
| 177 | Changjiang | 1,535 | 953 | 582 | 0.23 |
| 178 | Zhoushan | 1,474 | 796 | 678 | 1.16 |
| 179 | Xinyu | 1,471 | 0 | 1,471 | 1.17 |
| 180 | Nyingchi | 1,448 | 260 | 1,188 | 0.20 |
| 181 | Weifang | 1,372 | 930 | 442 | 9.36 |
| 182 | Qianxinan | 1,371 | 514 | 857 | 2.84 |
| 183 | Baishan | 1,347 | 674 | 673 | 1.20 |
| 184 | Changji | 1,326 | 744 | 582 | 1.60 |
| 185 | Chongzuo | 1,203 | 777 | 426 | 2.07 |
| 186 | Changdu | 1,181 | 369 | 812 | 0.68 |
| 187 | Baoting | 1,168 | 460 | 708 | 0.17 |
| 188 | Hotan | 1,146 | 671 | 475 | 2.14 |
| 189 | Linfen | 1,118 | 793 | 325 | 4.46 |
| 190 | Tunchang County | 1,090 | 489 | 601 | 0.27 |
| 191 | Qitaihe | 1,087 | 569 | 518 | 0.87 |
| 192 | Fuxin | 1,065 | 823 | 242 | 1.78 |
| 193 | Zhangzhou | 980 | 335 | 645 | 5.05 |
| 194 | Yulin4 | 967 | 461 | 506 | 5.76 |
| 195 | Shihezi | 945 | 802 | 143 | 0.60 |
| 196 | Matsubara | 930 | 330 | 600 | 2.78 |
| 197 | Jixi | 923 | 553 | 370 | 1.84 |
| 198 | Qinzhou | 902 | 491 | 411 | 3.24 |
| 199 | Haibei | 900 | 577 | 323 | 0.28 |
| 200 | Tongren | 893 | 893 | 0 | 3.14 |
| 201 | Dingan County | 882 | 494 | 388 | 0.29 |
| 202 | Altay | 824 | 446 | 378 | 0.62 |
| 203 | Wuzhishan | 806 | 429 | 377 | 0.11 |
| 204 | Ganzi | 779 | 192 | 587 | 1.18 |
| 205 | Karamay | 760 | 392 | 368 | 0.42 |
| 206 | Chaoyang | 750 | 704 | 46 | 2.95 |
| 207 | Baise | 722 | 402 | 320 | 3.62 |
| 208 | Nujiang | 720 | 377 | 343 | 0.54 |
| 209 | Aral | 711 | 365 | 346 | 0.33 |
| 210 | Tower | 705 | 481 | 224 | 1.35 |
| 211 | Wuzhong | 705 | 429 | 276 | 1.39 |
| 212 | Yingkou | 704 | 348 | 356 | 2.44 |
| 213 | Ningde | 690 | 446 | 244 | 2.89 |
| 214 | Shizuishan | 672 | 481 | 191 | 0.80 |
| 215 | Ordos | 630 | 458 | 172 | 2.06 |
| 216 | Ximeng | 629 | 458 | 171 | 1.00 |
| 217 | Shuangyashan | 609 | 185 | 424 | 1.46 |
| 218 | Leshan | 585 | 313 | 272 | 3.27 |
| 219 | Hainan | 585 | 253 | 332 | 0.48 |
| 220 | Baiyin | 583 | 262 | 321 | 1.72 |
| 221 | Chaozhou | 570 | 230 | 340 | 2.65 |
| 222 | Haixi | 566 | 458 | 108 | 0.52 |
| 223 | Chifeng | 552 | 487 | 65 | 4.31 |
| 224 | Yanbian | 522 | 379 | 143 | 2.10 |
| 225 | Yanan | 520 | 492 | 28 | 2.25 |
| 226 | Liaoyuan | 512 | 352 | 160 | 1.18 |
| 227 | Wenshan | 500 | 282 | 218 | 3.62 |
| 228 | Yili | 496 | 419 | 77 | 4.62 |
| 229 | Shannan | 494 | 212 | 282 | 0.34 |
| 230 | Rizhao | 485 | 326 | 159 | 2.90 |
| 231 | Maoming | 480 | 172 | 308 | 6.12 |
| 232 | Qiongzong | 479 | 287 | 192 | 0.23 |
| 233 | Guigang | 475 | 261 | 214 | 4.33 |
| 234 | Shuozhou | 455 | 249 | 206 | 1.77 |
| 235 | Baisha | 451 | 262 | 189 | 0.12 |
| 236 | Xian | 450 | 450 | 0 | 8.83 |
| 237 | Meishan | 446 | 219 | 227 | 3.00 |
| 238 | Xingan League | 439 | 91 | 348 | 1.60 |
| 239 | Wulanchabu | 434 | 332 | 102 | 2.11 |
| 240 | Bayannaoer | 423 | 275 | 148 | 1.68 |
| 241 | Mianyang | 398 | 288 | 110 | 4.81 |
| 242 | Shigatse | 397 | 288 | 109 | 0.72 |
| 243 | Alxa League | 389 | 286 | 103 | 0.25 |
| 244 | Aksu | 373 | 202 | 171 | 2.46 |
| 245 | Wuhai | 369 | 230 | 139 | 0.56 |
| 246 | Tongliao | 367 | 201 | 166 | 3.12 |
| 247 | Wujiaqu | 357 | 103 | 254 | 0.09 |
| 248 | Bazhou | 357 | 216 | 141 | 1.28 |
| 249 | Qiannan | 348 | 299 | 49 | 3.26 |
| 250 | Yichun | 332 | 29 | 303 | 1.10 |
| 251 | Ali | 326 | 178 | 148 | 0.10 |
| 252 | Zhongwei | 324 | 217 | 107 | 1.15 |
| 253 | Jiaxing | 321 | 45 | 276 | 4.61 |
| 254 | Zhengzhou | 319 | 83 | 236 | 9.72 |
| 255 | Huangnan | 318 | 142 | 176 | 0.27 |
| 256 | Kashgar | 309 | 177 | 132 | 4.21 |
| 257 | White | 306 | 253 | 53 | 1.91 |
| 258 | Cangzhou | 303 | 187 | 116 | 7.51 |
| 259 | Qingyang | 294 | 256 | 38 | 2.24 |
| 260 | Bijie | 265 | 227 | 38 | 6.64 |
| 261 | Anshun | 261 | 206 | 55 | 2.33 |
| 262 | Zibo | 241 | 134 | 107 | 4.69 |
| 263 | Jiuquan | 235 | 144 | 91 | 1.12 |
| 264 | Nagqu | 233 | 231 | 2 | 0.48 |
| 265 | Dingxi | 227 | 128 | 99 | 2.79 |
| 266 | Hechi | 220 | 107 | 113 | 3.50 |
| 267 | Chizhou | 214 | 191 | 23 | 1.44 |
| 268 | Tumshuk | 210 | 32 | 178 | 0.17 |
| 269 | Yangjiang | 204 | 96 | 108 | 2.53 |
| 270 | Jinchang | 203 | 147 | 56 | 0.47 |
| 271 | Liangshan | 199 | 84 | 115 | 4.82 |
| 272 | Turpan | 197 | 157 | 40 | 0.63 |
| 273 | Hulunbeir | 196 | 151 | 45 | 2.53 |
| 274 | Jinzhong | 187 | 18 | 169 | 3.35 |
| 275 | Yaan | 184 | 130 | 54 | 1.54 |
| 276 | Pingliang | 175 | 129 | 46 | 2.10 |
| 277 | Golow | 175 | 167 | 8 | 0.20 |
| 278 | Daxinganling | 158 | 45 | 113 | 0.44 |
| 279 | Yulin2 | 155 | 72 | 83 | 3.38 |
| 280 | Binzhou | 146 | 69 | 77 | 3.89 |
| 281 | Zhaoqing | 143 | 112 | 31 | 4.08 |
| 282 | Zhangye | 143 | 52 | 91 | 1.22 |
| 283 | Qiqihar | 143 | 85 | 58 | 5.05 |
| 284 | Linxia | 142 | 58 | 84 | 2.03 |
| 285 | Jiayuguan | 130 | 55 | 75 | 0.25 |
| 286 | Lishui | 127 | 41 | 86 | 2.17 |
| 287 | Suihua | 121 | 81 | 40 | 5.21 |
| 288 | Guyuan | 119 | 99 | 20 | 1.22 |
| 289 | Heyuan | 110 | 37 | 73 | 3.08 |
| 290 | Mudanjiang | 110 | 59 | 51 | 2.63 |
| 291 | Wuzhou | 108 | 61 | 47 | 3.02 |
| 292 | Kezhou | 107 | 11 | 96 | 0.62 |
| 293 | Luliang | 107 | 11 | 96 | 3.85 |
| 294 | Taiyuan | 103 | 0 | 103 | 4.34 |
| 295 | Tianshui | 101 | 82 | 19 | 3.32 |
| 296 | Heihe | 99 | 38 | 61 | 1.64 |
| 297 | Yushu | 94 | 87 | 7 | 0.41 |
| 298 | Baoji | 94 | 94 | 0 | 3.78 |
| 299 | Laiwu | 94 | 65 | 29 | 1.38 |
| 300 | Yunfu | 93 | 44 | 49 | 2.48 |
| 301 | Yingtian | 88 | 9 | 79 | 1.16 |
| 302 | Tongchuan | 81 | 60 | 21 | 0.85 |
| 303 | Pingxiang | 76 | 0 | 76 | 1.91 |
| 304 | Jiamusi | 76 | 38 | 38 | 2.36 |
| 305 | Shaoxing | 76 | 44 | 32 | 4.99 |
| 306 | Xinzhou | 72 | 19 | 53 | 3.16 |
| 307 | Shanwei | 70 | 43 | 27 | 3.04 |
| 308 | Dezhou | 68 | 24 | 44 | 5.79 |
| 309 | Jinhua | 63 | 0 | 63 | 5.52 |
| 310 | Meizhou | 61 | 41 | 20 | 4.36 |
| 311 | Hami | 61 | 31 | 30 | 0.61 |
| 312 | Lhasa | 60 | 60 | 0 | 0.60 |
| 313 | Yuncheng | 59 | 42 | 17 | 5.31 |
| 314 | Gannan | 51 | 26 | 25 | 0.71 |
| 315 | Liaocheng | 36 | 0 | 36 | 6.04 |
| 316 | Zhaotong | 35 | 35 | 0 | 5.48 |
| 317 | Jinan | 30 | 30 | 0 | 7.23 |
| 318 | Guangyuan | 28 | 19 | 9 | 2.64 |
| 319 | Hegang | 26 | 19 | 7 | 1.04 |
| 320 | Luoyang | 21 | 0 | 21 | 6.80 |
| 321 | Tongling | 18 | 0 | 18 | 1.60 |
| 322 | Chuzhou | 17 | 0 | 17 | 4.04 |
| 323 | Huzhou | 16 | 0 | 16 | 2.98 |
| 324 | Bozhou | 13 | 7 | 6 | 5.10 |
| 325 | Taian | 11 | 0 | 11 | 5.64 |
| 326 | Quzhou | 10 | 0 | 10 | 2.16 |
| 327 | Huaibei | 10 | 0 | 10 | 2.21 |
| 328 | Zaozhuang | 9 | 0 | 9 | 3.92 |
| 329 | Huaihua | 8 | 0 | 8 | 4.92 |
| 330 | Bengbu | 7 | 0 | 7 | 3.33 |
| 331 | Huainan | 7 | 0 | 7 | 3.46 |
| 332 | Xuancheng | 6 | 0 | 6 | 2.60 |
| 333 | Hengshui | 6 | 0 | 6 | 4.45 |
| 334 | Longnan | 6 | 0 | 6 | 2.60 |
| 335 | Hefei | 0 | 0 | 0 | 7.87 |
| 336 | Ganzhou | 0 | 0 | 0 | 8.59 |
| 337 | Shuanghe | 0 | 0 | 0 | 0.05 |
| 338 | Maanshan | 0 | 0 | 0 | 2.78 |
| 339 | Bazhou | 0 | 0 | 0 | 0.94 |
| 340 | Linyi | 0 | 0 | 0 | 10.44 |
| 341 | Beitun | 0 | 0 | 0 | 0.08 |
| 342 | Yibin | 0 | 0 | 0 | 4.51 |
| 343 | Shangqiu | 0 | 0 | 0 | 7.28 |
| 344 | Taizhou4 | 0 | 0 | 0 | 4.65 |
| 345 | Shaoyang | 0 | 0 | 0 | 7.32 |
| 346 | Heze | 0 | 0 | 0 | 8.62 |
| 347 | Yichun | 0 | 0 | 0 | 5.53 |
| 348 | Wuxi | 0 | 0 | 0 | 6.53 |
| 349 | Fuyang | 0 | 0 | 0 | 7.99 |
| 350 | Yutian County,<br>Xinjiang | 0 | 0 | 0 | 0.22 |
| 351 | Xuzhou | 0 | 0 | 0 | 8.71 |
| 352 | Suqian | 0 | 0 | 0 | 4.88 |
| 353 | Hetian County,<br>Xinjiang | 0 | 0 | 0 | 0.28 |
| 354 | Huaian | 0 | 0 | 0 | 4.89 |
| 355 | Kaifeng | 0 | 0 | 0 | 4.55 |
| 356 | Nanjing | 0 | 0 | 0 | 8.27 |
| 357 | Loudi | 0 | 0 | 0 | 3.89 |
| 358 | Suzhou4 | 0 | 0 | 0 | 5.60 |
| 359 | Macau | 0 | 0 | 0 | 0.63 |
| 360 | Jining | 0 | 0 | 0 | 8.35 |
| 361 | Qiandongnan | 0 | 0 | 0 | 3.51 |
| 362 | Kokodala | 0 | 0 | 0 | 0.08 |
| 363 | Xianyang | 0 | 0 | 0 | 4.99 |
| 364 | Lianyungang | 0 | 0 | 0 | 4.50 |
| 365 | Gejiu, Yunnan | 0 | 0 | 0 | 0.47 |
| 366 | Shangrao | 0 | 0 | 0 | 6.75 |
| 367 | Moyu County, Xinjiang | 0 | 0 | 0 | 0.53 |
| 368 | Wuhu | 0 | 0 | 0 | 3.67 |
| 369 | Sanmenxia | 0 | 0 | 0 | 2.26 |
| *Data derived from user geolocation data from Tencent (https://heat.qq.com). Cities are sorted according to the overall travel volume to and from Wuhan. These data also are available from github (https://github.com/ZhanweiDU/2019nCov.git). |  |  |  |  |  |

## Notes

### Competing Interest Statement

The authors have declared no competing interest.

## References

1. Wuhan Municipal Health Commission. Wuhan Municipal Health Commission briefing on the pneumonia epidemic situation 31 Dec 2019 [In Chinese] [Internet]. 2020 [cited 2020 Jan 11]. http://wjw.wuhan.gov.cn/front/web/showDetail/2019123108989

2. Statement on the second meeting of the International Health Regulations (2005) Emergency Committee regarding the outbreak of novel coronavirus (2019-nCoV) [Internet]. [cited 2020 Feb 5]. Available from: https://www.who.int/news-room/detail/30-01-2020-statement-on-the-second-meeting-of-the-international-health-regulations-(2005)-emergency-committee-regarding-the-outbreak-of-novel-coronavirus-(2019-nCoV)

3. Imai N, Dorigatti I, Cori A, Donnelly C, Riley S, Ferguson NM. MRC Centre for Global Infectious Disease Analysis. News 2019-nCoV. Report 2: Estimating the potential total number of novel coronavirus cases in Wuhan City, China. [cited 05 Feb 2020]. https://www.imperial.ac.uk/media/imperial-college/medicine/sph/ide/gida-fellowships/2019-nCoV-outbreak-report-22-01-2020.pdf

4. Enserink M. War stories. Science. 2013;339:1264–8. PubMed https://doi.org/10.1126/science.339.6125.1264

5. World Health Organization. Disease outbreak news: novel coronavirus–Thailand (ex-China) [Internet] 14 Jan 2020 [cited 2020 Jan 27]. https://www.who.int/csr/don/14-january-2020-novel-coronavirus-thailand-ex-china/en

6. Wilder-Smith A, Teleman MD, Heng BH, Earnest A, Ling AE, Leo YS. Asymptomatic SARS coronavirus infection among healthcare workers, Singapore. Emerg Infect Dis. 2005;11:1142–5. PubMed https://doi.org/10.3201/eid1107.041165

7. MOBS Lab. 2019 nCOV [Internet]. [cited 2020 Jan 26]. https://www.mobs-lab.org/2019ncov.html

8. Li Q, Guan X, Wu P, Wang X, Zhou L, Tong Y, et al. Early transmission dynamics in Wuhan, China, of novel coronavirus–infected pneumonia. N Engl J Med. 2020 Jan 29 [Epub ahead of print] PubMed https://doi.org/10.1056/NEJMoa2001316

9. Statista. Most popular messaging apps as of October 2019, based on the number of monthly active users. [cited 2020 Jan 31]. https://www.statista.com/statistics/258749/most-popular-global-mobile-messenger-apps

10. Chan JF-W, Yuan S, Kok K-H, To KK-W, Chu H, Yang J, et al. A familial cluster of pneumonia associated with the 2019 novel coronavirus indicating person-to-person transmission: a study of a family cluster. Lancet. 2020 Jan 24 [Epub ahead of print]. PubMed https://doi.org/10.1016/S0140-6736(20)30154-9

11. Wuhan Municipal Health Commission. Bulletin on pneumonitis associated with new coronavirus infection [In Chinese] [Internet]. [cited 2020 Jan 29]. http://wjw.wuhan.gov.cn/front/web/list2nd/no/710

## References

1. Bogoch II, Watts A, Thomas-Bachli A, Huber C, Kraemer MUG, Khan K. Pneumonia of unknown etiology in Wuhan, China: potential for international spread via commercial air travel. J Travel Med. 2020 Jan 14 [Epub ahead of print]. PubMed https://doi.org/10.1093/jtm/taaa008

2. Wu JT, Leung K, Leung GM. Nowcasting and forecasting the potential domestic and international spread of the 2019-nCoV outbreak originating in Wuhan, China: a modelling study. Lancet (2020) https://doi.org/10.1016/S0140-6736(20)30260-9

3. Lai S, Bogoch II, Watts A, Khan K, Li Z, Tatem A. Preliminary risk analysis of 2019 novel coronavirus spread within and beyond China. https://www.worldpop.org/resources/docs/china/WorldPop-coronavirus-spread-risk-analysis-v1-25Jan.pdf

4. Li Q, Guan X, Wu P, Wang X, Zhou L, Tong Y, et al. Early transmission dynamics in Wuhan, China, of novel coronavirus–infected pneumonia. N Engl J Med. 2020 Jan 29 [Epub ahead of print] PubMed https://doi.org/10.1056/NEJMoa2001316

5. Huang C, Wang Y, Li X, Ren L, Zhao J, Hu Y, et al. Clinical features of patients infected with 2019 novel coronavirus in Wuhan, China. Lancet. 2020 Jan 24 [Epub ahead of print]. PubMed https://doi.org/10.1016/S0140-6736(20)30183-5

6. Imai N, Dorigatti I, Cori A, Donnelly C, Riley S, Ferguson NM. MRC Centre for Global Infectious Disease Analysis. News 2019-nCoV. Report 2: estimating the potential total number of novel coronavirus cases in Wuhan City, China. London: Imperial College London; 21 Jan 2020. [cited 2020 Jan 26]. https://www.imperial.ac.uk/media/imperial-college/medicine/sph/ide/gida-fellowships/2019-nCoV-outbreak-report-22-01-2020.pdf

7. Cauchemez S, Fraser C, Van Kerkhove MD, Donnelly CA, Riley S, Rambaut A, et al. Middle East respiratory syndrome coronavirus: quantification of the extent of the epidemic, surveillance biases, and transmissibility. Lancet Infect Dis. 2014;14:50–6. PubMed https://doi.org/10.1016/S1473-3099(13)70304-9

8. Donnelly CA, Ghani AC, Leung GM, Hedley AJ, Fraser C, Riley S, et al. Epidemiological determinants of spread of causal agent of severe acute respiratory syndrome in Hong Kong. Lancet. 2003;361:1761–6. PubMed https://doi.org/10.1016/S0140-6736(03)13410-1

9. Backer JA, Klinkenberg D, Wallinga J. The incubation period of 2019-nCoV infections among travellers from Wuhan, China. [medRxiv preprint of Infectious Diseases (except HIV/AIDS) January 28, 2020]. https://doi.org/10.1101/2020.01.27.20018986

10. Hay J. Turning nCoV case reports into infection incidence [cited 2020 Jan 31]. Github. https://github.com/jameshay218/case_to_infection

11. Lauer SA, Grantz KH, Bi Q, Jones FK, Zheng Q, Meredith H, et al. The incubation period of 2019-nCoV from publicly reported confirmed cases: estimation and application. medRxiv. February 04, 2020https://doi.org/10.1101/2020.02.02.20020016

12. World Health Organization. Disease outbreak news: novel coronavirus–Thailand (ex-China) [Internet] 14 Jan 2020 [cited 2020 Jan 27]. https://www.who.int/csr/don/14-january-2020-novel-coronavirus-thailand-ex-china/en

13. Ministry of Health, Labour and Welfare, China. Development of pneumonia associated with the new coronavirus [in Chinese]. [cited 27 Jan 2020]. https://www.mhlw.go.jp/stf/newpage_08906.html

14. Gutierrez B, Hill S, Kraemer M, Loskill A, Mekaru S, Pigott D, et al. Epidemiological and demographic data of confirmed cases in the 2019-nCoV outbreak. Github [cited 2020 Jan 31]. https://github.com/BoXu123/2019-nCoV-epiData

15. MOBS Lab. 2019 nCOV [Internet]. [cited 31 Jan 2020]. https://www.mobs-lab.org/2019ncov.html

16. Models of Infectious Disease Agent Study (MIDAS). Central resource of data and information in support of modeling research on the 2019 novel coronavirus (2019-nCoV). Google Docs [cited 2020 Feb 2]. https://docs.google.com/document/d/1pL6ogED0Qix08V0zjJbbNVhf6yf-xBShWVxLbXKVLXI/edit

17. Genomic epidemiology of novel coronavirus (nCoV). [cited 2020 Feb 2]. https://nextstrain.org/ncov

18. Wang L, Wu JT. Characterizing the dynamics underlying global spread of epidemics. Nat Commun. 2018;9:218. PubMed https://doi.org/10.1038/s41467-017-02344-z

19. Scalia Tomba G, Wallinga J. A simple explanation for the low impact of border control as a countermeasure to the spread of an infectious disease. Math Biosci. 2008;214:70–2. PubMed https://doi.org/10.1016/j.mbs.2008.02.009

20. Gautreau A, Barrat A, Barthélemy M. Global disease spread: statistics and estimation of arrival times. J Theor Biol. 2008;251:509–22. PubMed https://doi.org/10.1016/j.jtbi.2007.12.001

21. Chinese Center for Disease Control and Prevention. Epidemic update and risk assessment of 2019 novel coronavirus [cited 31 Jan 2020]. http://www.chinacdc.cn/yyrdgz/202001/P020200128523354919292.pdf

22. Real-time surveillance of pneumonia epidemics in China [Internet]. [cited 2020 Jan 27]. https://3g.dxy.cn/newh5/view/pneumonia?scene=2&clicktime=1579579384&enterid=1579579384&from=timeline&isappinstalled=0

